# Evaluation of the efficacy of *Ashwagandha* in chronic knee pain among older adults with knee osteoarthritis – Protocol of a double-blind randomized controlled trial

**DOI:** 10.64898/2026.06.23.26356318

**Authors:** Pallavi Mundada, Vaishali Kuchewar, Manoj Raut, Disha Garg, Bharti

## Abstract

**Background:** Chronic knee pain, primarily due to knee osteoarthritis, significantly impairs quality of life of the elderly by restricting mobility, reducing physical activity, and contributing to psychological distress such as depression and anxiety. The standard of care include pharmacological treatments (e.g., NSAIDs, analgesics), physical therapy, lifestyle modifications, and, in severe cases, surgical interventions, most of which often do not provide sustained relief, may carry adverse effects, and lead to poly-pharmacy, particularly in elderly patients with comorbidities. *Ashwagandha* is known to have *Balya* (strengthening) and *Rasayana* (adaptogenic/ rejuvenating) properties, pacifies *Vata* and thus may be helpful in mitigating chronic musculoskeletal pain.

**Objective:** To evaluate the efficacy of oral *Ashwagandha* & *Til Taila Abhyanga* for six weeks on chronic knee pain, functionality, mobility, quality of life, general wellbeing, sleep quality of the older adults with knee osteoarthritis.

**Materials & Methods:** Patients of any gender, above 60 years age and having pain in one or both knee joints since more than 3 months and average severity rated ≥4 on the Wong-Baker Faces, due to knee osteoarthritis diagnosed as per the American College of Rheumatology Criteria are being included in the study. Medically unstable and non-ambulatory patients with severity Grade>4 of Kellgren and Lawrence scale for OA, BMI≥30 kg/m^2^, those on recent treatment with intra-articular injections or Ayurveda medications, having knee implant or fixed flexion deformity in knees, history of acute trauma, or with severe systemic/infectious ailments or other chronic conditions affecting the knee joint are excluded.

Total 72 participants are enrolled and allocated randomly to either group. Participants are given either *Ahwagandha Churna* or *Boswellia* extract as oral medication for 45 days. *Til Taila* and standard operating procedure of *Abhyanga* (external oleation through massage) at the affected knee are given in both groups. The severity of pain is assessed by the numeric pain rating scale after every 15 days. Other outcomes are change in Knee Injury and Osteoarthritis Outcome Score, score of WHO Wellbeing Index -5, Global Sleep Assessment Questionnaire, Five Times Sit and Stand test and Time to Up and Go, after the intervention period. The need for conventional analgesics through the study duration is also observed and will be compared in both groups.

**Discussion:** The outcomes of this double-blind randomized controlled trial will inform about the efficacy of *Ashwagandha* which is generally considered as *Balya* in alleviating chronic pain of knee osteoarthritis among elderly. This study can generate evidence and lead to larger effectiveness studies on role of adding *Ashwagandha* to the standard care for management of chronic musculoskeletal pain in older adults.

## BACKGROUND

Chronic musculoskeletal pain, as defined by the International Classification of Diseases 11th Revision (ICD-11), encompasses persistent pain that occurs in muscles, bones, or joints, lasting for more than three months and often resulting from underlying pathological conditions such as osteoarthritis (OA).[1] Among older adults, chronic knee pain, primarily due to knee osteoarthritis, significantly impairs quality of life by restricting mobility, reducing physical activity, and contributing to psychological distress such as depression and anxiety.[2]

Current management strategies for chronic knee pain include pharmacological treatments (e.g., NSAIDs, analgesics), physical therapy, lifestyle modifications, and, in severe cases, surgical interventions like knee arthroplasty.[3] However, these modalities often fall short in providing sustained relief and may carry adverse effects, particularly in elderly patients with comorbidities.[4] This highlights the necessity for more effective, holistic, and safer treatment alternatives.

Ayurveda, the ancient Indian system of medicine, offers promising approaches for managing musculoskeletal and arthritic pain.[5] Various *Guggulu* (*Commiphora mukul*) formulations (like *Yogaraj Guggulu*) and herbo-mineral formulations (like *Vatavidhvamsa Rasa*) are commonly prescribed to manage the pain in knee osteoarthritis. Multi-herb decoctions and hydro-alcoholic extracts (like *Rasnasaptak Kwath* and *Dashamoolarishta*) and medicated oils (like *Narayan Taila* etc.) are frequently bought even over the counter by the patients suffering from arthritic pain in India.[6]

However, for long-term use in chronic painful musculoskeletal conditions, availability and cost-effectiveness of multiple Ayurveda medicines can be a concern especially for the geriatric age group. The issues like multi-morbidity, poly-pharmacy, dependency and isolation are common among the older adults. So therapeutic interventions including lesser number of medications that can comprehensively benefit the health through improving the vitality and relieve the symptoms; that can be consumed or used without any assistance at home by the patients and, that can be cost-effective are important approaches that should be focussed in the geriatric health care.

### Rationale

Single herb medications with multi-dimensional efficacy in context with the health in geriatric age group have much scope of research in this scenario. *Ashwagandha* (*Withania somnifera*), a cornerstone herb, is renowned for its adaptogenic, anti-inflammatory and analgesic properties.[7] It has been documented to enhance physical function and reduce pain in various musculoskeletal conditions, supporting its benefit for elderly patients.[8] Further, when combined with a simple but effective procedure like *Abhyanga* (therapeutic message with oil) on the affected joint can be a sustainable option to the use of conventional analgesics like NSAIDs. *Abhyanga* with various oils (sesame, mustard, castor, etc. or oils after processing to extract active compounds from medicinal herbs) has been traditionally acclaimed for its ability to alleviate pain, enhance joint function, and improve overall well-being. However, no RCTs on the combination of oral *Ashwagandha* and *Til Taila Abhyanga* in the selected population are available. Further, though *Ashwagandha* is known to act as a *Balya* (strengthening)*, Brimhaniya* (nourishing) and *Rasayana* (adaptogenic/ rejuvenating) medicine, [9,10] its role in pain management among elderly with chronic musculoskeletal pain is not compared with *Shallaki* (*Boswellia*) extract (a routinely prescribed Ayurvedic and herbal analgesic for arthritis) is not known.

So, this clinical trial aims to determine whether the combination of oral *Ashwagandha* and *Til Taila Abhyanga* for 6 weeks is as effective as oral *Shallaki* and *Til Taila Abhyanga* on chronic knee pain and associated disability among the older adults (above 60 years age) with knee osteoarthritis

## OBJECTIVES

The primary objective of this study is to evaluate the efficacy of *Ashwagandha Churna* as compared to *Boswellia* extract on chronic knee pain perceived by the older adults with knee osteoarthritis. The secondary objectives are to compare the effect of the interventions on functionality, mobility, quality of life, general wellbeing, sleep quality of the elderly participants.

## METHODS

### Patient and public involvement

The patients were not involved in designing of the trial however, as the study is a clinical trial, it is conducted involving the patients attending the Outpatient department of (OPD) of the Central Ayurveda Research Institute, Delhi.

### Trial design

This is an interventional parallel group with equal allocation non-inferiority double-blind randomized controlled clinical trial.

### Study Setting

OPD of an Institute dedicated for research in Ayurveda at Delhi in India.

### Eligibility

Patients with chronic knee pain since ≥ 3 months duration and of severity in the past week ≥4 on Wong-Baker Faces [11,12], and who fulfill the clinical American College of Rheumatology (ACR) criteria for knee osteoarthritis.

### Inclusion Criteria

Patients of any gender, above 60 years age and having pain in one or both knee joints since more than 3 months with average severity of knee pain in the past week is reported ≥4 when assessed through Wong-Baker Faces, fulfilling the clinical American College of Rheumatology (ACR) criteria for diagnosis of knee osteoarthritis i.e., pain should accompany with at least three of the following: age >50 years, stiffness <30 min, crepitus, bony tenderness, bony enlargement and no palpable warmth,[13,14,15] are included. The participants are also screened for having likely unimpaired cognition by including the patients scoring ≥3 in the Mini-Cog scale,[16] to be included in the study.

### Exclusion Criteria

Non-ambulatory and medically unstable patients presenting with abnormal blood pressure (≥160/100 or ≤90/68 mm Hg)[17], blood sugar level (greater than 180 mg/dL or below 70 mg/dL)[18,19] or those showing signs of severe osteoarthritis on radiological evaluation and eligible to be categorized as Grade-4 of Kellgren and Lawrence scale for OA,[20] unilateral/bilateral fixed flexion deformity in knees or metallic implant in knee joint; or severe deformity of lower limbs (e.g., knee varus or knee valgus) or those with BMI≥30 kg/m², history of treatment with intra-articular injection of corticosteroids or hyaluronic acid or oral corticosteroids within a period of three months preceding study, knee joint replacement, arthroscopy within preceding one year or other surgery on lower limbs within the past 6 months or significant acute trauma; or other systemic ailments like unstable angina, arrhythmia, severe vision problems, or neurological dysfunction or mental illness on medication, myocardial infarction or cerebrovascular accident (CVA) in the past 6 months or other, or on regular intake of Ayurveda medicines (including any Ayurveda procedures) or physiotherapy (including therapeutic exercises) for pain relief in the past 2 weeks are excluded. Known cases of rheumatoid arthritis, inflammatory [e.g., systemic lupus erythematosus (SLE), psoriatic arthritis], or infective arthritis, crystal arthritis (gout or pseudo-gout) or polyarthralgia, or hemophilic arthropathy, or bone and soft tissue tumours or any other active malignancy or substance use disorder (CAGE score≥2)[21], any other surgical or medical condition including acute/sub-acute infection or prescription (like patients on sedatives) or any such factor that limits the patient’s participation in the study or jeopardizes the outcomes or those with known history of hypersensitivity to the trial intervention or its constituents were also not included in the study.

## INTERVENTIONS

The participants in Group-1 receive a bottle with capsules containing *Ashwagandha* (*Withania somnifera*) root powder to be consumed in a dose of 3 gm/day i.e., two capsules thrice daily and the participants in Group-2 receive a bottle with capsules containing *Shallaki* (*Boswellia serrata*) extract to be consumed in a dose of 1.2 gm/day i.e., two capsules thrice daily. The participants in both arms also receive one or two bottles containing 75 ml sesame oil (depending upon number of knees involved) for topical application in the form of *Abhyanga* (Ayurvedic massage/ external oleation through massage with specific oil)[22] as per the prescribed standard operating procedure (SoP) twice daily. The intervention period is 45 days. Details are given in Table No. 1. The medicines are provided at every follow-up scheduled fortnightly. The SoP of *Abhyanga* is explained and provided to the study participants in print and audio-visual form. It includes the following steps - pouring oils over the affected knee joint and then gently massaging/ rubbing with light pressure in circular direction (clockwise and anti-clockwise) at all the surfaces of the knee for minimum 5 minutes so that the prescribed dose of 2.5 ml oil is appropriately absorbed in each affected knee joint.[23] The medicines are procured from a GMP certified reputed pharmacy and have been checked for quality as per available standards.

**Table No. 1:**
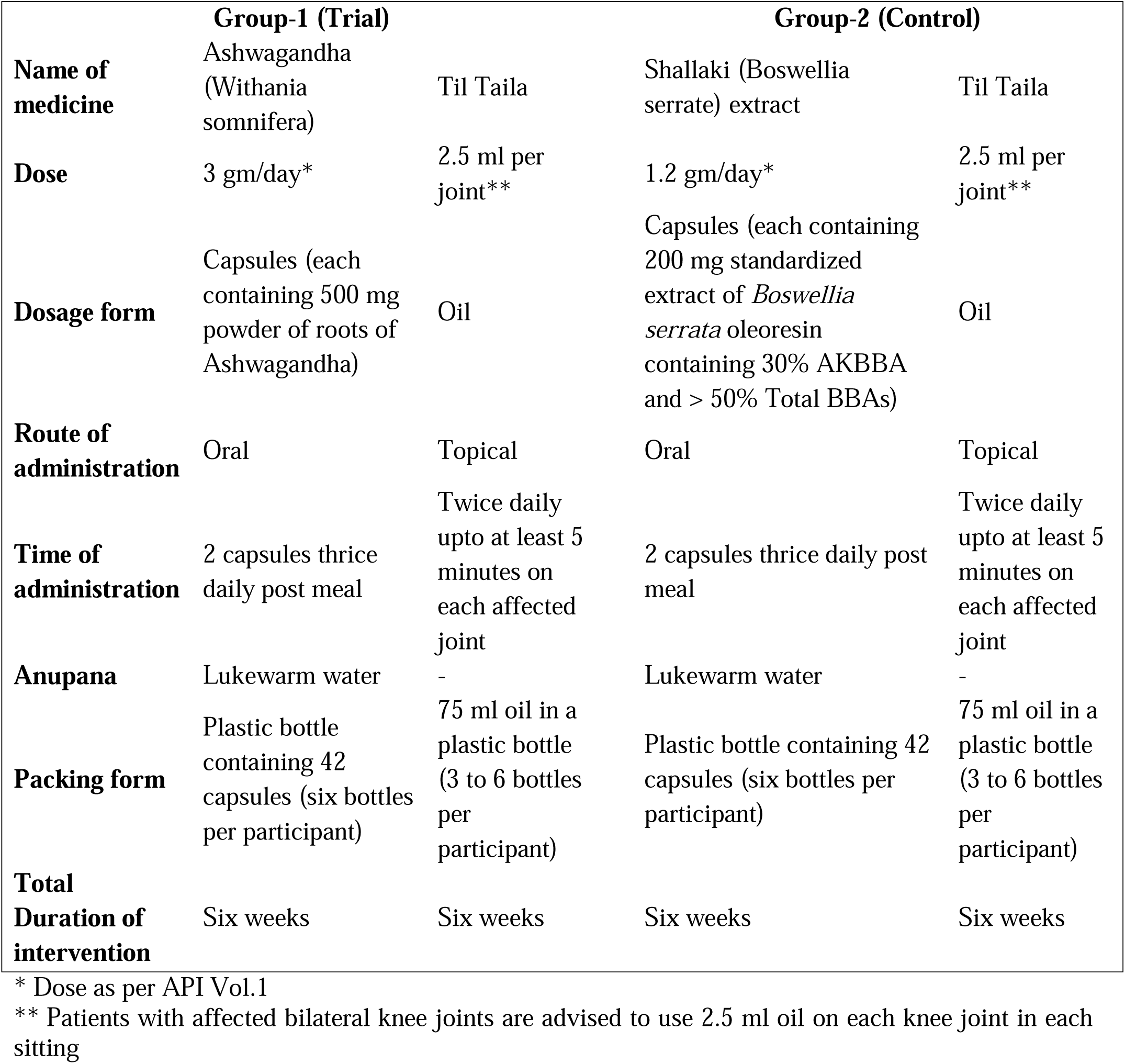
Details of trial intervention and comparator.

### Procedure for monitoring adherence

A compliance form is at the baseline, 15^th^ day and 30^th^ day follow-up visit to the study participants to tick after the medicine is consumed and *Abhyanga* is done each time in a day. The form is collected from them at every follow-up visit. The empty bottles of medicines are also collected for assessment of compliance.

### Adherence improvement strategy

The participants are called on telephone on the 7^rd^, 21^st^ day and 35^th^ day for reminding them about the medications and follow-up visits. Any requirement of rescue medicines or any adverse event is also enquired and noted in the CRF.

### Relevant concomitant medications permitted and prohibited

The participants reporting ongoing medication for any co-morbidity at baseline are advised to continue their treatment. Any other medicine if taken for symptoms of knee osteoarthritis is noted as rescue medication in the case record forms. Ancillary care in the form of Ayurveda medicines viz., *Punarnavadi Mandoor*, *Nisha Amalki, Laghu Sutshekha Rasa* and *Swadishta Virechan Churna* for anemia, diabetes, hyperacidity and constipation respectively are prescribed on the OPD paper for a short duration if required.

### Withdrawal Criteria

The participants not willing to continue in the trial and/or not adhering to treatment (compliance < 80%) are withdrawn from the study. Incidence of any condition mentioned in the exclusion criteria during the study period is also a criterion for withdrawal.

## OUTCOMES

Change in the score of Numeric Pain Rating Scale (NRS)[24] from the baseline is the primary outcome measure. The secondary outcome measures include Change in knee-related quality of life score, knee pain and symptom outcome score and Activities of Daily Living (ADL) score of the Knee Injury and Osteoarthritis Outcome Score (KOOS) questionnaire,[25,26] change in range of motion of the affected knee joint(s) measured by using a manual goniometer, change in Timed Up and Go test (TUG)[27] for measuring balance, change in Five-Times-Sit-to-Stand Test (FTSST)[28] for measuring muscle strength of the lower limbs, change in the score of WHO-5 Wellbeing Index score,[29,30] change in the Global Sleep Assessment Questionnaire (GSAQ) score,[31] and frequency of use of conventional analgesic medicines as rescue medication for symptomatic relief in knee pain during the study period, as compared to baseline in both groups. Incidence of treatment emergent adverse events (like skin irritation, weight gain, gastric reactions, changes in vitals like blood pressure etc.) will also be compared in both groups.

NRS is a scale with numbers marked from zero to ten wherein zero depicting no pain and ten indicating worst pain imaginable. The patient is asked to mark a number that shows her perception of worst pain during the past twenty-four hours. TUG assesses the length of time it takes participants to get up from a standard height chair, walk three meters, turn and return to the chair, and sit down again. The participant has to rise from a chair and return to a seated position as quickly as possible with their arms folded across their chests. The time to complete five repetitions is recorded for two separate trials, with a one min rest between each trial. The mean of the two trials is computed and used in the analysis.

### Participant timeline

The primary outcome NRS for knee pain is assessed at baseline and at every follow-up (biweekly) however, the other outcomes like KOOS, Goniometry, WHO-5 Wellbeing Index, GSAQ, TUG, FTSST are assessed at the baseline visit and at the last visit after six weeks of treatment. Assessment of treatment adherence and need of any rescue medication is done at every visit. The adverse events are investigated as soon as they are reported by the study participants. Table No. 2 contains detailed participant timeline.

**Table No. 2:**
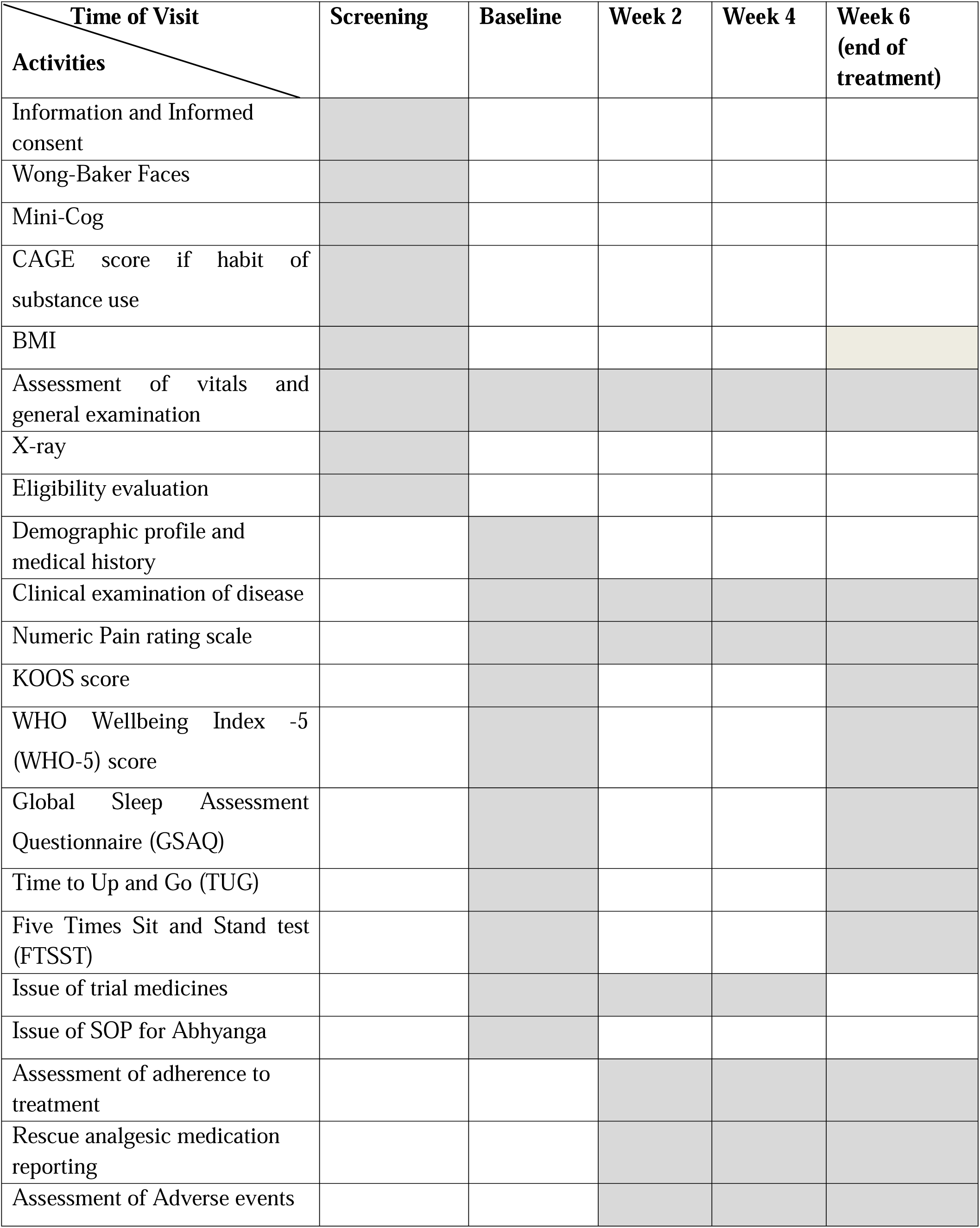
Participant Timeline.

### Sample size

Assuming the baseline mean (±SD) of the primary outcome i.e. Numeric Pain Rating Scale (NRS) to be 6.7 (±2.2) units based on a descriptive study on chronic pain among elderly[32] and a non-inferiority margin of 1.65 units which is the minimal clinically important difference (MCID) for NRS[33], the true/allowable difference in mean score of NRS in both groups is supposed to be 0, the sample size calculated (with the help of https://riskcalc.org/samplesize/) for 90% power and 5% significance level came out to be 62. Assuming a 15% attrition rate and rounding off, the total sample size was 72, which is to be equally allocated in both groups (36 in each group).

### Recruitment

All the physicians at the institutes are informed about the study and eligibility criteria. A flyer with brief information about the study is placed on the notice board in the geriatric OPD and the special OPD for joints. The target sample size is anticipated to be achieved in the defined timelines because, the footfall of elderly patients with complaints related to knee joint is sufficient at the study site.

### Sequence generation

Block randomization with unequal block sizes method is used. A computer-generated random sequence for the allocation of study participants in either group is created centrally in the Biostatistics department.

### Allocation concealment mechanism

The investigators as well as the participants are not aware of which group they are allocated to. The randomization list will be with the stats section until analysis.

### Blinding (masking)

The participants, care providers and outcome assessors are blinded in this study. The internal medicines will be administered through hard gelatin capsules of same size and smell. The bottles of capsules are labeled as per sequence numbers after randomization by the stats section. The figure no. 1 shows the similarity of bottles provided to the two groups. In case of occurrence of any serious treatment-emergent adverse event, un-blinding will be done and necessary action will be taken.

**Figure no. 1.**
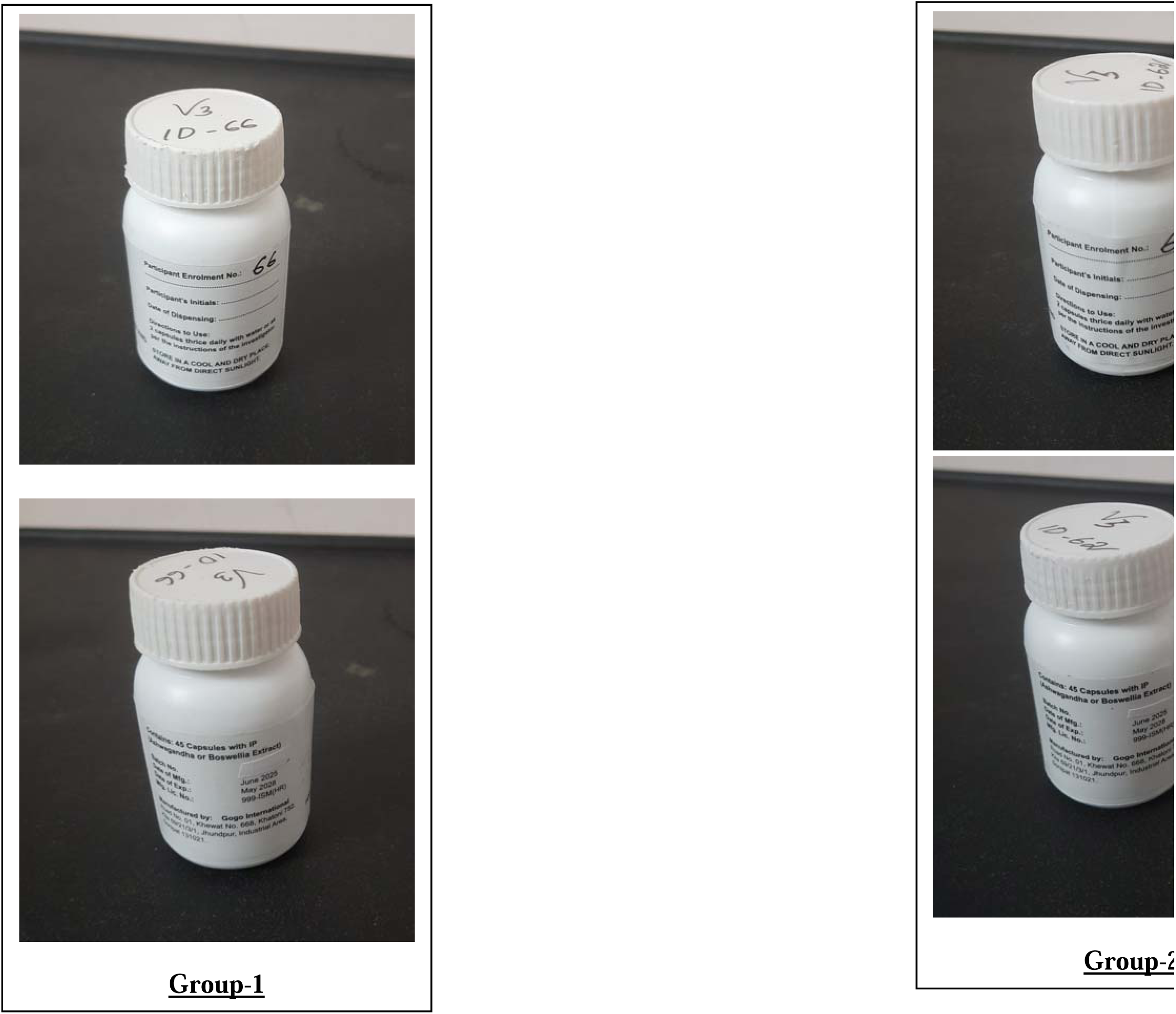
Similarity of bottles of the oral medicine given to the participants in two groups ensuring masking.

### Data collection methods

The eligible patients are informed about the study methods as per the participant information sheet (PIS) and those willing to participate are screened after their consent signing. They are clinically examined for general condition, vitals, BMI, and other criteria for inclusion and exclusion including radiological and bio-chemical evaluation. The data is collected in a case record form (CRF) prepared for entering findings at screening, baseline and follow-up visits as per Table No. 2. At the baseline visit, information regarding personal identification, demographic profile, detailed medical and surgical history and history of present illness is collected. The knee joints are examined and data related to outcome measures is collected. The Ayurvedic parameters like *Prakriti* (constitution)*, Agni* (various aspects of digestive capacity), possible *Hetu* (possible causative factors for vitiation of *Doshas*) are also assessed and recorded at the baseline. During the follow-up visits on day 15^th^ and day 30^th^, the status of chief complaints, NRS, compliance assessment, concomitant medicines, need for rescue medicines, adverse events, and clinical assessment are reported. At the final visit i.e. 45^th^ day, assessment of all outcome measures are done. The participants are called on telephone on regular intervals to ensure treatment adherence and their continuity in the trial. The adverse events are recorded as soon as reported by the participants during telephonic conversations or follow-up visits.

Attempt to contact the participants who discontinue is made to collect information on recent adverse event, reason of discontinuation and present status of pain in NRS along with need of rescue medications.

### Data management and monitoring

The data from the source documents are entered in an excel sheet in the form of codes for most of the variables and is meticulous inspected for quality. Periodic data cleaning is done to minimalize missing data. The source documents and Excel sheets containing participant’s data are kept confidential and safe. The data in the excel sheets will be anonymized and will be identified only through enrolment numbers after completion of the trial, before statistical analysis. The trial is conducted by the principal investigator. The study supervisor monitors the data collection methods and data quality through periodic surprise checks. The status reports and adverse events related reports are communicated to the ethics and scientific committees of the University and study site periodically.

### Statistical methods

The data of categorical variables will be presented as numbers (percentage) and will be compared between groups using the chi-square test, while within-group comparison will be done by using the McNemar’s/Cochrane Q-test. Continuous data will be checked for normality. The data at each follow-up will be compared with the baseline data within group using rANOVA tests and, between groups using independent sample t-test if it follows normal distribution. It will be presented as mean ± SD. The pre-post comparison will be made through paired sample t-test/Wilcoxon signed rank test. Non-normal data will be presented as median and interquartile range and will be compared within group using Friedman test and, between groups comparison will be done using Mann-Whitney U test. A p-value of <0.05 will be considered as significant.

Per protocol approach will be used for statistical analysis as this is a non-inferiority trial. Any instances of missing data will be handled by immediate cross-check with the source documents and appropriate imputation method will be applied if required.

## DISCUSSION

*Ashwagandha* (*Withania somnifera*) is a widely cultivated, not endangered herb. Classical texts generously describe its properties as *Vata* and *Kapha* dosha pacifying, beneficial in leukoderma, inflammatory and degenerative conditions, strengthening, rejuvenating and enhancing spermatogenesis.[34] The pharmacological studies on *Ashwagandha* prove its anti-inflammatory, antioxidant, and analgesic effects. The bioactive compounds in *Ashwagandha*, such as withanolides, contribute to its therapeutic effects by modulating inflammatory pathways and reducing oxidative stress.[35] Sumantran VN et al. have explored the chondro-protective potential of root (aqueous) extracts of *Withania somnifera* in osteoarthritis and described the mechanism of action in detail. A review explored the Clinical Trial Registry of India for the Ashwagandha related trials and found that 12 trials on musculoskeletal pain and bone-related problems were registered during the period of 2008 to 2020.[36] Ashwagandha has also been found effective in inflammatory arthritic conditions when used as a single intervention [37] and with a herbo-mineral preparation (*Siddha Makardhwaj*)[38]. A double blind RCT showed that Ashwagandha root extract 250 mg twice daily was effective in significantly reducing the symptoms of knee osteoarthritis. The authors of the above study attribute the analgesic activity of Ashwagandha mostly to withaferin A, which has been shown to reduce the production of prostaglandins, mediating pain by inhibiting cyclo-oxygenase (COX). They also suggest that the soothing action of Ashwagandha on the nervous system may contribute to the analgesic effects.[39] However evidence on the efficacy and feasibility of Ashwagandha root powder in improving the chronic knee pain in the geriatric age group is not available. So, this study is postulated to compare its effect on the older adults suffering with chronic knee pain due to osteoarthritis with that of *Boswellia* extract which is commonly prescribed as herbal analgesic for musculoskeletal pain. Local application of sesame oil as per the method prescribed by Ayurveda for *Abhyanga* is added to the intervention to maintain patient satisfaction as the study is conducted in India where oil application for joint pain is an over-the-counter medication practice. This minimised the possibility of the potential confounding effect of its use by some of the study participants.

## Data Availability

Not applicable as this is protocol article

## ETHICS AND DISSEMINATION

### Research ethics approval

The protocol is approved by the Institutional Ethics Committee of MGAC, DMIHER, Wardha on 17.3.2025 and the Central Ethics Committee at CARI, Punjabi Bagh, New Delhi on 14.5.2025.

### Protocol amendments

No amendment has been made in the study protocol since initiation to date.

### Consent or assent & Confidentiality

The study is being conducted as per the Declaration of Helsinki and ICMR ethics guidelines 2017. The consent material like participant information sheet and the consent form are approved by the Ethics Committees. The participants are screened only after obtaining their informed written consent. Anonymization of the data will be done before analysis and confidentiality regarding the medical records of the study participants is maintained.

### Access to data

The data will be available with the Investigator and the University (DMIHER).

### Ancillary and post-trial care

The participants are advised to seek the consultation from the physicians at the regular OPDs of the Institute as routine care after their participation in the trial.

### Dissemination policy

The results of the study will be updated in the Clinical Trial Registry of India (CTRI) portal where the trial is registered. The final report will also be published in a reputed journal and can be presented in scientific conferences.

### Appendices

Informed consent materials, Case record form, SPIRIT 2025 Checklist

### Trial Registration

This clinical trial is prospectively registered in CTRI on 11.6.2025 with number CTRI/2025/06/088610

